# Ultra-low-field MRI for imaging of severe multiple sclerosis: a case-controlled study

**DOI:** 10.1101/2025.08.22.25332332

**Authors:** Niels Bergsland, Alex Burnham, Michael G Dwyer, Alex Bartnik, Ferdinand Schweser, Cheryl Kennedy, Ashley Tranquille, Mehak Semy, Ella Schnee, David Young-Hong, Svetlana Eckert, David Hojnacki, Christine Reilly, Ralph HB Benedict, Bianca Weinstock-Guttman, Robert Zivadinov

**Affiliations:** Buffalo Neuroimaging Analysis Center, Department of Neurology, Jacobs School of Medicine and Biomedical Sciences, University at Buffalo, State University of New York, Buffalo, NY, USA; The Boston Home, Dorchester, MA, USA; Center for Biomedical Imaging, University at Buffalo, State University of New York, Buffalo, NY, USA; Department of Neurology, Jacobs Comprehensive MS Treatment and Research Center, Jacobs School of Medicine and Biomedical Sciences, University at Buffalo, State University of New York, Buffalo, NY, USA

**Author notes:** **Corresponding author:** Robert Zivadinov, MD, PhD, SUNY Distinguished Professor of Neurology, Director, Buffalo Neuroimaging Analysis Center, Director, Center for Biomedical Imaging at the Clinical Translational Science Institute, Department of Neurology, Jacobs School of Medicine and Biomedical Sciences, University at Buffalo, State University of New York, 77 Goodell Street, Suite 450, Buffalo, NY 14203.

**Keywords:** multiple sclerosis, severe MS, ultralow-field MRI, gray matter, thalamus, cortical, atrophy, disability

## Abstract

**Background:** Severe multiple sclerosis (MS) presents challenges for clinical research due to mobility constraints and specialized care needs. Traditional MRI studies often exclude this population, limiting understanding of severe MS progression. Portable, ultra-low-field MRI enables bedside imaging.

**Objectives:** To (i) assess the feasibility of portable MRI in severe MS, (ii) compare measurement approaches for automated tissue volumetry from ultra-low-field MRI.

**Methods:** This prospective study enrolled 40 progressive MS patients (24 severely disabled, 16 less severe) from academic and skilled nursing settings. Participants underwent 0.064T MRI for tissue volumetry using conventional and artificial intelligence (AI)-driven segmentation. Clinical assessments included physical disability and cognition. Group comparisons and MRI-clinical associations were assessed.

**Results:** MRI passed rigorous quality control, reflecting complete brain coverage and lack of motion artifact, in 38/40 participants. In terms of severe versus less severe disease, the largest effect sizes were obtained with conventionally-calculated gray matter (GM) volume (partial η^2^=0.360), cortical GM volume (partial η^2^=0.349), and whole brain volume (partial η^2^=0.290) while an AI-based approach yielded the highest effect size for white matter volume (partial η^2^=0.209). For clinical outcomes, the most consistent associations were found using conventional processing while AI-based methods were dependent on algorithm and input image, especially for cortical GM volume.

**Conclusion:** Portable, ultralow-field MRI is a feasible bedside tool that can provide insights into late-stage neurodegeneration in individuals living with severe MS. However, careful consideration is required in implementing tissue volumetry pipelines as findings are heavily dependent on the choice of algorithm and input.

## Introduction

A subset of people with multiple sclerosis (pwMS) experience a particularly severe form of the disease, which may be evident early after onset or develop later, following two disability trajectories.^1–4^ In early aggressive MS, individuals reach an Expanded Disability Status Scale (EDSS) score of ≥6.0 within 10 years—or before age 40—from disease onset,^5^ whereas those with late severe MS attain similar disability levels after more than 10 years of disease duration or after 40 years of age.^2–4^ Early aggressive MS typically results in rapid and marked disability, with an estimated prevalence of 5–10% among pwMS.^1, 5^ Late severe MS progresses more gradually, likely being influenced by aging-related processes.^2–4, 6, 7^ Some individuals with severe MS eventually develop an even more advanced state of disability, marked by severe ambulatory impairment (EDSS ≥7.0) and/or substantial cognitive decline. These pwMS often require wheelchair or bed confinement, assistance with daily activities, psychosocial support, rehabilitation, and professional care; they represent about 30% of those with severe MS.^2^

Little research has been conducted to characterize severe MS, in part because there are many practical difficulties in studying this population.^2–4^ Those living with severe MS are often confined to their own homes or skilled nursing facilities, where constant, intensive care is available.^2–4^ The high-dependency nature of their conditions means that transporting them to academic research facilities for participation in studies is not only a logistical challenge but also a potential health risk. The need for specialized transport and caregivers to accompany them may lead to prohibitive costs and overall infeasibility. In fact, many pwMS with a severe form of the illness do not obtain MRI or other examinations for monitoring disease activity once they progress to this stage. Furthermore, clinical and academic study settings are generally unequipped to manage the complex needs of these individuals, who may require medical equipment, emergency care, or additional accommodations, increasing the difficulty of conducting longitudinal studies.

In the Comprehensive Analysis of Severely Affected Multiple Sclerosis (CASA-MS) phase 1 study, we showed significantly lower cortical gray matter (GM) and thalamic volume in severe compared to less severe MS.^4^ Moreover, no significant differences between the two groups were observed in terms of lesion volume, suggesting that GM atrophy may be more relevant for development of severe disability. A major weakness of that retrospective study was that MRI data was collected from different local MRI facilities for those with a severe phenotype, leading to uncontrolled heterogeneity in MRI outcomes while the less severe group was acquired on the same 3T scanner. This was inevitable as ambulatory and cognitive disabilities rendered it logistically infeasible to transport them to specialized MRI centers.

Portable, ultralow-field MRI technology presents a promising opportunity for studying the vulnerable population of individuals with severe MS, as it offers a means to safely and effectively gather imaging data right at their bedside, minimizing distress and maximizing comfort.^8, 9^ Ultra-low-field MRI (e.g., 64 millitesla [0.064T]) systems have been cleared by the Food and Drug Administration and offer greater accessibility and cost efficiency compared to traditional scanners.^8, 10, 11^ However, ultra-low-field images are also of comparatively lower quality, with reduced signal-to-noise ratios and spatial resolution compared to standard MRI scanners. To overcome this limitation, deep learning techniques have been developed to improve image quality and facilitate volumetric segmentation from ultralow-field MRI acquisitions.^10, 12–15^

To address the limitation of heterogeneous MRI acquisitions in the CASA-MS phase 1 study, we assessed the feasibility of using standardized, ultra-low-field MRI to image fully disabled individuals with the goal of comparing different tissue volumetry approaches with respect to previously reported group differences. Based on our preliminary findings, we hypothesized that individuals with severe MS would be characterized by evidence of lower GM volume compared to their less severe counterparts.

## Materials and Methods

### Study population

The CASA-MS phase 2 was a prospective, cross-sectional study that was conducted at The Boston Home (TBH), a specialized residential MS facility in Dorchester, Massachusetts and at a tertiary MS center at the University at Buffalo (UB), Buffalo, New York. The study enrolled 24 severely disabled pwMS from TBH. The study inclusion criteria were: 1) age between 30-80 years old, 2) diagnosed with MS based on the 2017-revised McDonald criteria,^16^ 3) having progressive MS, according to the 2013 Lublin criteria,^17^ and 4) having either early disability accrual severe MS (n=12, <10 years of disease duration or <40 years of age at the time of reaching EDSS of ≥6.0) or late disability accrual severe MS (n=12, ≥10 years of disease duration or ≥40 years of age at the time of reaching EDSS of ≥6.0). For comparison purposes, 16 age-, sex-, and disease duration matched progressive pwMS from UB with less severe disease were also enrolled. The exclusion criteria were: 1) having experienced a relapse or having received steroids within 30 days prior to study enrollment or MRI examination, 2) women who were pregnant or lactating, and 3) inability to obtain informed consent from the study participant or their medical proxy.

Informed consent was obtained from all participants, in accordance with the protocols approved by the UB Institutional Review Board. In instances where participants were unable to consent due to physical or cognitive limitations, a legally authorized representative provided consent on their behalf.

### Clinical assessments

A standardized questionnaire was used to collect demographic and clinical information. In addition, study participants underwent clinical assessments by trained medical doctors, with oversight from a board-certified neurologist (BWG).

Physical disability was assessed as previously reported.^2^ Specifically, we utilized the EDSS,^18^ timed 25-foot walk test (T25FWT),^19^ 9-hole peg test (9HPT) ^20^ and Scripps Neurological Rating Scale (SNRS).^21^ Four measures (EDSS, SNRS, T25FWT and 9HPT) allowed for calculation of the Combinatorial Weight-Adjusted Disability Score (CombiWISE).^22^ A maximum of 180 seconds was scored in the event of a study participant being unable to walk or taking longer time to complete the T25FWT. Similarly, a maximum of 300 seconds was allowed for the 9HPT. Lower SNRS and higher Combi-WISE scores indicate greater disability.

All neuropsychological tests were performed by trained investigators under supervision of board-certified neuropsychologist (RHBB). Cognitive function was assessed as previously published,^3^ by using Auditory Test of Processing Speed (ATOPS)^23^ and Brief International Cognitive Assessment of MS (BICAMS)^24^ test batteries. For the purpose of exploring the association between ultra-low-field MRI and cognition, we only used ATOPS and Symbol Digit Modalities Test (SDMT), ^25^ given that both measure processing speed – one of the most commonly affected cognitive area in MS.^26^

### MRI acquisition and analyses

The brains of all pwMS were scanned on the same ultralow-field 0.064T MRI scanner (Hyperfine Swoop, Guilford, CT, USA) that was securely transported between the two cities in a truck and scanner manufacturer-provided cradle. The imaging protocol was set up to match that of previously published studies using the same scanner model in people with MS,^15, 27^ which included axial and sagittal T1 weighted imaging (WI), axial T2-WI, and axial and sagittal T2 fluid-attenuated inversion recovery (T2-FLAIR) sequences. The resolutions were 1.5 x 1.5 x 5 mm^3^ (T1-WI and T2-WI) and 1.6 x 1.6 x 5 mm^3^ (T2-FLAIR), respectively. Detailed scanning parameters can be found elsewhere.^15, 27^ The protocol was 35 min long. We developed a technique at the Boston Home using a Hoyer lift to move fully disabled study participants (weighing between 110 pounds/49.9 kg and 265 pounds/120.2 kg) from wheelchair/bed to the MRI table, followed by adequate positioning for scanning (assisted by 3 trained operators) (Figure 1). We used padding and supports to help patients maintain a comfortable position. Low acquisition noise allowed the operators to stand next to the scanner and talk to the participant as needed during scanning or play music, if desired.

**Figure 1.**
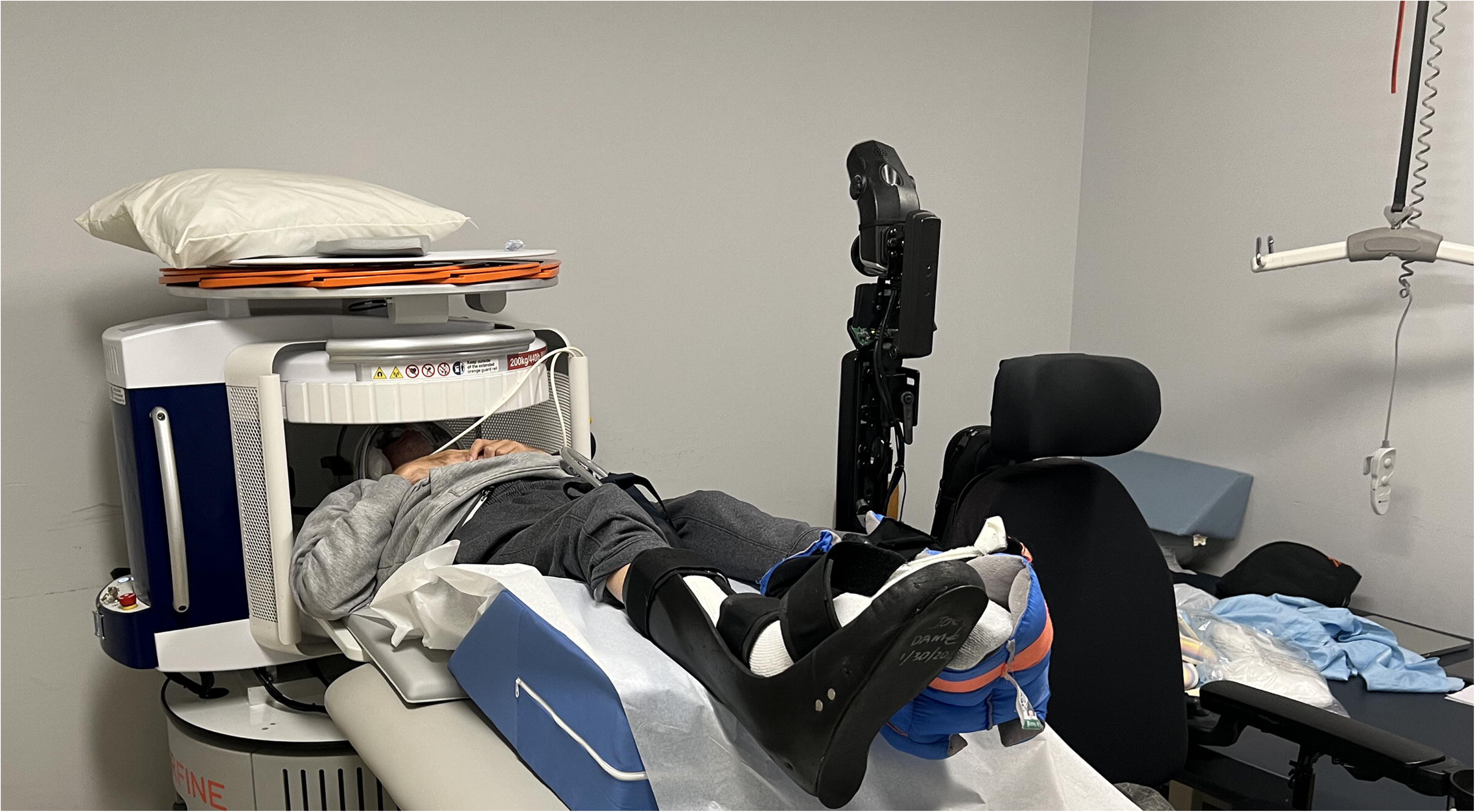
An FDA-cleared Hyperfine Swoop MRI ultralow-field MRI (64mT), allowing for imaging of people with severe multiple sclerosis at the bedside, is shown. The Hoyer lift can be seen on the right of the image, which was used to safely move the individual from his wheelchair to the MRI table and facilitate proper positioning into the head coil.

The image analysis was performed in a blinded manner without knowledge of the demographic details, clinical conditions and originating center. All images were subjected to visual inspection. Quality metrics assessed included overall quality indicators such as anatomical coverage, the presence of imaging artifacts, the extent of patient movement, noise levels, and image contrast.^28^

T2 lesion volume (LV) and T1-LV were obtained using a semi-automated contouring/thresholding technique.^29^ For regional tissue segmentation, we utilized three different approaches. First, regional volumes were determined on lesion filled^30^ axial T1-WI using Structural Image Evaluation using Normalisation of Atrophy (SIENAX, https://fsl.fmrib.ox.ac.uk/fsl/docs/#/structural/siena/index).^31^ Initial experiments resulted in poor brain extraction with SIENAX despite optimization of deskulling parameters. As such, we replaced the default BET tool with SynthStrip (https://surfer.nmr.mgh.harvard.edu/docs/synthstrip/)^32^ but kept BET for generating the skull mask. Second, we processed axial T1-WI images using the recon-all-clinical pipeline (https://surfer.nmr.mgh.harvard.edu/fswiki/recon-all-clinical)^33^, part of FreeSurfer version 7.4.1, which utilizes a combination of 1) SynthSeg^13^ to obtain a volumetric segmentation and linear registration to Talairach space; 2) SynthSR^34^ to obtain a higher resolution 1mm MPRAGE for visualization; 3) SynthDist to fit surfaces by predicting the distance maps and reconstructing topologically accurate cortical surfaces. Third, we processed axial T1-WI images using the WMH-SynthSeg tool (https://surfer.nmr.mgh.harvard.edu/fswiki/WMH-SynthSeg)^35^, which handle images of any contrast and resolution while providing lesion segmentations. The tool is specifically trained to handle images with low resolution and signal-to-noise, as is the case with ultra-low-field MRI. To assess replicability of results using different image contrasts, the recon-all-clinical and WMH-SynthSeg pipelines were also run using the axial T2-FLAIR images.

Inpainting was not used prior to the recon-all-clinical pipeline or WMH-SynthSeg as they both explicitly handle the presence of white matter (WM) lesions. From all three approaches, we obtained separate measures of whole brain (WB), GM, cortical GM, WM, and lateral ventricular volumes. Thalamic volume was obtained only from the recon-all-clinical and WMH-SynthSeg pipelines. Absolute volumes were obtained from each pipeline.

### Data availability

The data that support the findings of this study are available upon reasonable request to the corresponding author.

### Statistical analyses

All statistical analyses and visualizations were performed using SPSS version 28.0 (IBM, Armonk, NY, USA) and RStudio version 2023.09.1+494 (R Foundation for Statistical Computing, Vienna, Austria). Data distributions were assessed using visual inspection of the data and histograms, and model assumptions were checked using Q-Q plots. Categorical variables were compared using chi-square tests. Group comparisons were performed parametrically using Student’s t-test (for normally distributed data) and non-parametrically using Mann Whitney U-test. Being non-normally distributed, ATOPS underwent log transformation.

Analysis of covariance (ANCOVA), adjusted for age, sex, and either head size (SIENAX) or intracranial volume (recon-all-clinical and WMH-SynthSeg), was performed to assess differences in imaging measures between severe and less severe MS as well as between severe MS with early and delayed accrual of disability. Controlling for head size/intracranial volume has been shown to be more powerful compared to directly utilizing fractional values (e.g., brain volume divided by intracranial volume) in statistical analyses.^36^ Partial η^2^ was calculated as a measure of effect size, with 0.01, 0.06, and > 0.14 indicating small, medium, and large effects, respectively.

Partial correlations, adjusted for the same covariates as in ANCOVA, were used to assess relationships between clinical and MRI outcomes. Non-parametric Spearman correlations were used for EDSS while Pearson correlations were utilized for the other clinical outcomes.

The Benjamini-Hochberg procedure was used to adjust for multiple comparisons by controlling the false discovery rate (FDR) when comparing differences in imaging measures between groups and when correlating imaging measures with a given clinical outcome.

Corrected p-values lower than 0.05 were considered statistically significant.

Bland-Altman plots were created to assess the agreement between different post-processing techniques for automated tissue volumetry. Plots were inspected to detect systematic bias (i.e., consistent over- or under-estimates) as well as assess outliers or trends in disagreement across the range of values.

## Results

### Demographic and clinical characteristics of the study populations

Table 1 shows demographic and clinical characteristics in pwMS with severe and less severe disease, as well as when characterized by early or late accrual of disability.

**Table 1.** Demographic and clinical characteristics of the study population.

| **Demographic and clinical characteristics** | **Less severe MS**<br>**(n=16)** | **Severe MS**<br>**(n=24)** | **p-value** | **Early disability accrual severe MS**<br>**(n=12)** | **Late disability accrual severe MS**<br>**(n=12)** | **p-value** |
| --- | --- | --- | --- | --- | --- | --- |
| Age in years, mean (SD) | 60.6 (11.0) | 62.0 (9.5) | 0.673 | 62.2 (11.7) | 61.8 (7.3) | 0.918 |
| Age at onset in years, mean (SD) | 32.4 (10.9) | 31.7 (10.3) | 0.830 | 34.3 (13.3) | 29.1 (5.5) | 0.226 |
| BMI, mean (SD) | 26.7 (4) | 26.2 (4.7) | 0.712 | 26.6 (5.1) | 25.8 (4.4) | 0.672 |
| Education, years, mean (SD) | 14.5 (2.7) | 14.7 (2.9) | 0.724 | 14.7 (3.6) | 14.6 (2.6) | 0.474 |
| Disease duration (years), mean (SD) | 30.1 (10.7) | 30.3 (8.4) | 0.962 | 27.9 (9.4) | 32.7 (6.9) | 0.087 |
| Female, n (%) | 15 (52.5) | 8 (50) | 0.522 | 9 (75) | 6 (50) | 0.400 |
| Race, n (%)<br>White<br>African-American | 15 (93.8)<br>1 (6.2) | 23 (95.8)<br>1 (4.2) | 0.999 | 12 (100)<br>0 (0) | 11 (91.7)<br>1 (8.3) | 0.999 |
| Marital status, n (%)<br>Single<br>Married<br>Divorced/separated<br>Widowed | 2 (12.5)<br>12 (75)<br>0 (0)<br>2 (12.5) | 8 (33.3)<br>4 (16.7)<br>10 (41.7)<br>2 (8.3) | **<0.001** | 2 (16.7)<br>2 (25)<br>5 (41.7)<br>2 (16.7) | 6 (50)<br>1 (8.3)<br>5 (41.7)<br>0 (0) | 0.172 |
| Disease course, n (%)<br>Relapsing-progressive<br>Non-relapsing progressive<br>Primary-progressive | 9 (56.3)<br>6 (37.5)<br>1 (6.3) | 7 (29.2)<br>15 (62.5)<br>2 (8.3) | 0.228 | 5 (41.7)<br>7 (58.3)<br>0 (0) | 2 (16.7)<br>8 (66.7)<br>2 (16.7) | 0.187 |
| Comorbidities, n (%)<br>Hypertension<br>Hypercholesterolemia<br>Heart disease<br>Diabetes<br>Obesity<br>Smoking | 10 (62.5)<br>10 (62.5)<br>7 (43.8)<br>2 (12.5)<br>3 (18.8)<br>7 (43.8) | 7 (29.2)<br>10 (41.7)<br>4 (16.7)<br>7 (29.2)<br>6 (25)<br>9 (37.5) | 0.053<br>0.148<br>0.08<br>0.395<br>0.717<br>0.947 | 4 (33.3)<br>4 (33.3)<br>1 (8.3)<br>4 (33.3)<br>4 (33.3)<br>4 (33.3) | 3 (25)<br>6 (50)<br>3 (25)<br>3 (25)<br>2 (16.7)<br>5 (41.7) | 0.999<br>0.680<br>0.590<br>0.640<br>0.999<br>0.999 |
| Relapses in the last 24 months, mean | 0.2 (0.5) | 0.05 (0.2) | 0.332 | 0.09 (0.3) | 0 (0) | 0.205 |
| (SD) |  |  |  |  |  |  |
| EDSS, median (IQR) | 4.0 (3.1-6.0) | 8.0 (8.0-8.5) | **p<0.001** | 8.3 (8-8.5) | 8.0 (7.5-8.5) | 0.121 |
| SNRS, mean (SD) | 71.8 (16.1) | 36.1 (11.5) | **p<0.001** | 32.8 (10.5) | 39.3 (12.1) | 0.086 |
| CombiWISE, mean (SD) | 33.6 (12.7) | 78 (6.5) | **p<0.001** | 79.9 (5.8) | 76.2 (6.8) | 0.088 |
| SDMT, mean (SD) | 49.6 (12.3) | 23.5 (10.4) | **p<0.001** | 19.1 (10.6) | 27.4 (9) | 0.083 |
| ATOPS (log transformed), mean (SD) | 1.5 (0.1) | 1.7 (0.1) | **p<0.001** | 1.7 (0.2) | 1.7 (0.2) | 0.431 |
| DMT, n (%) |  |  |  |  |  |  |
| Interferon-β | 1 (6.3) | 0 (0) | **0.002** | 0 (0) | 0 (0) | 0.217 |
| Glatiramer acetate | 2 (12.5) | 3 (12.5) |  | 3 (25) | 0 (0) |  |
| Oral | 3 (18.8) | 0 (0) |  | 0 (0) | 0 (0) |  |
| Anti-CD20 | 3 (18.8) | 0 (0) |  | 0 (0) | 0 (0) |  |
| Natalizumab | 3 (18.8)) | 0 (0) |  | 0 (0) | 0 (0) |  |
| Other | 1 (6.3) | 0 (0) |  | 0 (0) | 0 (0) |  |
| No therapy | 3 (18.8) | 21 (87.5) |  | 0 (0) | 0 (0) |  |

**Legend:** MS – multiple sclerosis; SD-standard deviation; BMI-body mass index; EDSS-Expanded Disability Status Scale; IQR-interquartile range; SNRS-Scripts Neurological Rating Scale; Combi-WISE-Combinatorial Weight-Adjusted Disability Score; SDMT- Symbol Digit Modalities Test; ATOPS-Auditory Test of Processing Speed; DMT-disease-modifying treatment.
P-values derived from chi-square test, Student's t-test, Mann-Whitney U test and binomial negative regression. In bold are shown significant p values <0.05 and in italic trend p values <0.1.

A larger percentage of pwMS with severe disease completed ATOPS (23, 95.8%) compared to SDMT (19, 79.2%) (p=0.190). As expected, participants with severe MS had increased disability, as evidenced on all clinical assessments, including EDSS, CombiWISE, SNRS, SDMT and ATOPS (all p<0.001). The prevalence of cardiovascular comorbidities was similar between the two groups, except for a trend for higher prevalence of hypertension in those with less severe MS (p=0.053). Study participants with more severe MS had been single or divorced (p<0.001) and were not on any DMT (p=0.002) compared to their less severe counterparts.

Demographic and clinical characteristics between early versus late disability accrual were similar, except for numerically worse clinical outcomes in the early group for SNRS, Combi-WISE and ATOPS (all p<0.088).

### Feasibility of ultralow-field MRI

We were able to acquire ultra-low-field MRI images with whole brain coverage and minimal motion artifact in 38 out of 40 participants. Two individuals with severe MS were unable to be properly positioned within the head coil due to either having a very short neck or patient-reported discomfort, resulting in incomplete brain coverage.

### MRI differences between the study groups

Table 2 shows comparison of MRI outcomes between pwMS split by severe versus less severe disease, and split by early versus delayed accrual of disability.

**Table 2.** Comparison of volumetrics between the study groups.

| MRI-based measures | Less severe MS (n=16) | Severe MS (n=22)% | Partial $\eta^2$ | p-value | Early disability accrual severe MS (n=10)% | Late disability accrual severe MS (n=12) | Partial $\eta^2$ | p-value |
| --- | --- | --- | --- | --- | --- | --- | --- | --- |
| T2-lesion volume |  |  |  |  |  |  |  |  |
| Semi-automated | 23.8 (20.9) | 27.5 (15.9) | 0.013 | 0.545 | 22.5 (13.1) | 31.6 (17.3) | 0.083 | 0.993 |
| WMH-SynthSeg (FL) | 17.3 (8.3) | 18.8 (7.3) | 0.014 | 0.545 | 16.5 (7.5) | 20.8 (6.8) | 0.099 | 0.993 |
| T1-lesion volume |  |  |  |  |  |  |  |  |
| Semi-automated | 12.2 (13.8) | 12.3 (8.4) | < 0.001 | 0.952 | 10.2 (7.5) | 14.1 (9.0) | 0.075 | 0.993 |
| WMH-SynthSeg (T1) | 14.6 (12.3) | 16.0 (8.7) | 0.006 | 0.678 | 14.4 (8.0) | 17.3 (9.3) | 0.041 | 0.998 |
| Whole brain volume |  |  |  |  |  |  |  |  |
| SIENAX | 978.5 (104.2) | 847.4 (83.2) | 0.292 | **0.009** | 801.1 (86.0) | 886.0 (60.0) | 0.190 | 0.993 |
| recon-all-clinical (T1) | 886.7 (101.9) | 771.1 (126.4) | 0.245 | **0.017** | 700.4 (120.2) | 830.0 (101.6) | 0.013 | 0.998 |
| recon-all-clinical (FL) | 778.1 (131.6) | 704.8 (141.6) | 0.100 | 0.101 | 635.7 (140.4) | 762.3 (119.2) | 0.056 | 0.993 |
| WMH-SynthSeg (T1) | 855.8 (100.3) | 775.1 (82.7) | 0.218 | **0.025** | 734.6 (66.5) | 808.9 (81.9) | 0.016 | 0.998 |
| WMH-SynthSeg (FL) | 895.6 (107.3) | 826.3 (92.9) | 0.191 | **0.030** | 792.2 (83.4) | 854.7 (94.1) | < 0.001 | 0.998 |
| Gray matter volume |  |  |  |  |  |  |  |  |
| SIENAX | 479.7 (52.0) | 399.8 (44.5) | 0.360 | **0.003** | 384.4 (47.4) | 412.7 (39.3) | 0.097 | 0.993 |
| recon-all-clinical (T1) | 501.0 (53.8) | 437.0 (74.7) | 0.145 | **0.050** | 397.6 (76.3) | 470.0 (57.3) | 0.021 | 0.998 |
| recon-all-clinical (FL) | 468.0<br>(92.6) | 418.0<br>(97.1) | 0.066 | 0.182 | 370.0<br>(92.6) | 458.0<br>(84.5) | 0.057 | 0.993 |
| WMH-SynthSeg (T1) | 525.5<br>(60.3) | 481.6<br>(46.0) | 0.209 | **0.025** | 459.1<br>(42.2) | 500.3<br>(41.8) | 0.035 | 0.998 |
| WMH-SynthSeg (FL) | 571.6<br>(69.9) | 533.2<br>(55.0) | 0.173 | **0.039** | 511.6<br>(52.1) | 551.3<br>(52.6) | 0.001 | 0.998 |
| Cortical gray matter volume |  |  |  |  |  |  |  |  |
| SIENAX | 361.5<br>(40.6) | 307.5<br>(28.0) | 0.349 | **0.003** | 296.6<br>(29.4) | 316.6<br>(24.4) | 0.099 | 0.993 |
| recon-all-clinical (T1) | 367.9<br>(42.0) | 334.0<br>(44.5) | 0.037 | 0.331 | 306.4<br>(44.6) | 357.0<br>(29.6) | 0.028 | 0.998 |
| recon-all-clinical (FL) | 376.9<br>(59.4) | 341.0<br>(67.4) | 0.083 | 0.132 | 308.2<br>(68.9) | 368.3<br>(54.7) | 0.056 | 0.993 |
| WMH-SynthSeg (T1) | 404.0<br>(50.8) | 380.4<br>(36.7) | 0.025 | 0.404 | 364.1<br>(31.9) | 393.9<br>(36.0) | 0.009 | 0.998 |
| WMH-SynthSeg (FL) | 441.0<br>(57.9) | 421.8<br>(48.4) | 0.013 | 0.545 | 405.1<br>(45.0) | 435.7<br>(48.4) | < 0.001 | 0.998 |
| Thalamic volume<sup>#</sup> |  |  |  |  |  |  |  |  |
| recon-all-clinical (T1) | 10.7<br>(1.7) | 8.3<br>(2.1) | 0.260 | **0.025** | 7.7<br>(2.7) | 8.6<br>(1.7) | < 0.001 | 0.998 |
| recon-all-clinical (FL) | 9.8<br>(2.2) | 7.6<br>(2.5) | 0.207 | **0.040** | 7.7<br>(2.2) | 7.6<br>(2.7) | 0.007 | 0.998 |
| WMH-SynthSeg (T1) | 11.0<br>(1.9) | 9.5<br>(1.4) | 0.192 | **0.040** | 8.8<br>(1.2) | 9.7<br>(1.5) | < 0.001 | 0.998 |
| WMH-SynthSeg (FL) | 11.1<br>(2.0) | 9.8<br>(1.6) | 0.110 | 0.110 | 9.3<br>(1.1) | 10.1<br>(1.9) | 0.006 | 0.998 |
| White matter volume |  |  |  |  |  |  |  |  |
| SIENAX | 498.8<br>(56.0) | 447.6<br>(49.3) | 0.137 | 0.054 | 416.7<br>(41.5) | 473.3<br>(40.5) | 0.251 | 0.957 |
| recon-all-clinical (T1) | 385.7<br>(49.8) | 334.1<br>(56.1) | 0.209 | **0.024** | 302.8<br>(48.4) | 360.1<br>(49.5) | < 0.001 | 0.998 |
| recon-all-clinical (FL) | 310.1<br>(45.7) | 286.9<br>(51.4) | 0.056 | 0.218 | 265.8<br>(52.7) | 304.4<br>(45.0) | 0.002 | 0.998 |
| WMH-SynthSeg (T1) | 330.2<br>(41.3) | 293.5<br>(40.2) | 0.165 | **0.040** | 275.5<br>(30.5) | 308.6<br>(42.2) | 0.003 | 0.998 |
| WMH-SynthSeg (FL) | 323.9<br>(39.1) | 293.1<br>(41.7) | 0.151 | **0.046** | 280.6<br>(34.9) | 303.4<br>(45.4) | 0.002 | 0.998 |
| Ventricular volume |  |  |  |  |  |  |  |  |
| SIENAX | 55.1<br>(21.1) | 68.7<br>(24.4) | 0.200 | **0.026** | 61.4<br>(24.9) | 74.8<br>(23.2) | 0.014 | 0.998 |
| recon-all-clinical (T1) | 43.4<br>(23.5) | 51.6<br>(27.5) | 0.074 | 0.157 | 39.8<br>(61.4) | 61.4<br>(27.9) | 0.068 | 0.993 |
| recon-all-clinical (FL) | 33.2<br>(20.2) | 44.4<br>(30.0) | 0.105 | 0.101 | 38.6<br>(27.7) | 48.7<br>(32.1) | 0.003 | 0.998 |
| WMH-SynthSeg (T1) | 51.7<br>(22.8) | 64.2<br>(28.8) | 0.120 | 0.078 | 53.6<br>(25.5) | 73.1<br>(29.5) | 0.025 | 0.998 |
| WMH-SynthSeg (FL) | 46.0<br>(20.7) | 59.0<br>(32.0) | 0.104 | 0.101 | 50.4<br>(28.4) | 66.2<br>(34.2) | 0.006 | 0.998 |

**Legend:** MS – multiple sclerosis; SD – standard deviation; T1 – T1-weighted; FL – T2-weighted
weighted fluid attenuated inversion recovery
% - Two individuals with early disability accrual in the severe MS group was excluded due to incomplete coverage of the brain

We did not find significant differences in T2-LV and T1-LV between groups, regardless of whether a semi-automated or a fully automated, AI-based segmentation approach (i.e., WMH-SynthSeg) was utilized. In terms of comparing those with severe versus less severe disease, the largest effect sizes for GM volume (partial η^2^=0.360), cortical GM volume (partial η^2^=0.349), and WB volume (partial η^2^=0.292), and lateral ventricular volume (partial η^2^=0.200) were found with SIENAX (all p ≤ 0.026), while WM volume (partial η^2^=0.209) and thalamic volume were found significant with recon-all-clinical using the T1W image (partial η^2^=0.260) (p = 0.024 and p=0.025, respectively). No significant differences were seen when comparing the early disability *vs.* late disability accrual groups, regardless of which method was used, although a large effect size was seen for WM volume with SIENAX (partial η^2^=0.251).

### Relationship between clinical outcomes and MRI measures

Table 3 shows partial correlation analyses adjusted for age and sex between clinical outcomes and MRI measures in the study population. Neither T2-LV nor T1-LV measures showed significant associations with any of the assessed disability measures. All SIENAX-derived measurements were significantly related to all clinical outcomes, except for WM volume with respect to SNRS and ATOPS. In terms of the two AI-driven approaches, recon-all-clinical and WMH-SynthSeg did not consistently yield the same results, even when the same image was used for processing. This was particularly notable for the thalamus, where the recon-all-clinical volume obtained from the T1W image was significantly associated with all clinical outcomes, whereas this was only the case for SDMT and ATOPS when using WMH-SynthSeg. Moreover, for a given approach, correlations were often discrepant with respect to the input image (i.e., T1W or FLAIR) used for processing. Unlike with SIENAX, WM volumes obtained from the recon-all-clinical and WMH-SynthSeg pipelines were significantly correlated with all clinical outcomes except for SNRS when using WMH-SynthSeg data from the FLAIR image. Regardless of image type and AI-driven pipeline, cortical GM volume was not significantly associated with EDSS.

**Table 3.** Relationship between clinical outcomes and MRI measures in the total study population.

| **MRI-based measures** | **EDSS** | **SNRS** | **Combi-WISE** | **SDMT** | **ATOPS** |
| --- | --- | --- | --- | --- | --- |
| T2-lesion volume |  |  |  |  |  |
| Semi-automated | 0.146<br>(0.433) | -0.170<br>(0.343) | 0.145<br>(0.444) | -0.256<br>(0.185) | 0.208<br>(0.262) |
| WMH-SynthSeg (FL) | 0.198<br>(0.291) | -0.210<br>(0.283) | 0.117<br>(0.558) | -0.265<br>(0.185) | 0.155<br>(0.422) |
| T1-lesion volume |  |  |  |  |  |
| Semi-automated | 0.082<br>(0.653) | -0.077<br>(0.655) | 0.045<br>(0.799) | -0.181<br>(0.368) | 0.126<br>(0.485) |
| WMH-SynthSeg (T1) | 0.200<br>(0.291) | -0.129<br>(0.498) | 0.081<br>(0.679) | -0.273<br>(0.179) | 0.185<br>(0.342) |
| Whole brain volume |  |  |  |  |  |
| SIENAX | **-0.672**<br>(**< 0.001**) | **0.451**<br>(**0.026**) | **-0.547**<br>(**0.009**) | **0.498**<br>(**0.001**) | **-0.507**<br>(**0.008**) |
| recon-all-clinical (T1) | **-0.485**<br>(**0.012**) | **0.500**<br>(**0.020**) | **-0.507**<br>(**0.013**) | **0.662**<br>(**< 0.001**) | **-0.585**<br>(**0.004**) |
| recon-all-clinical (FL) | **-0.403**<br>(**0.044**) | 0.326<br>(0.092) | -0.333<br>(0.084) | 0.108<br>(0.598) | **-0.437**<br>(**0.021**) |
| WMH-SynthSeg (T1) | **-0.489**<br>(**0.012**) | **0.481**<br>(**0.020**) | **-0.498**<br>(**0.014**) | **0.664**<br>(**< 0.001**) | **-0.526**<br>(**0.005**) |
| WMH-SynthSeg (FL) | -0.368<br>(0.058) | **0.461**<br>(**0.024**) | **-0.471**<br>(**0.018**) | **0.529**<br>(**0.006**) | **-0.459**<br>(**0.014**) |
| Gray matter volume |  |  |  |  |  |
| SIENAX | **-0.722**<br>(**< 0.001**) | **0.503**<br>(**0.020**) | **-0.602**<br>(**0.005**) | **0.529**<br>(**0.006**) | **-0.570**<br>(**0.004**) |
| recon-all-clinical (T1) | -0.372<br>(0.058) | **0.437**<br>(**0.027**) | **-0.423**<br>(**0.029**) | **0.537**<br>(**0.002**) | **-0.456**<br>(**0.015**) |
| recon-all-clinical (FL) | **-0.420**<br>(**0.039**) | 0.277<br>(0.147) | -0.286<br>(0.133) | 0.009<br>(0.961) | -0.349<br>(0.062) |
| WMH-SynthSeg (T1) | **-0.485** | **0.490** | **-0.504** | **0.618** | **-0.468** |
|  | **(0.012)** | **(0.028)** | **(0.013)** | **(0.001)** | **(0.013)** |
| WMH-SynthSeg (FL) | **-0.390**<br>**(0.044)** | **0.428**<br>**(0.028)** | **-0.441**<br>**(0.029)** | **0.429**<br>**(0.031)** | -0.339<br>(0.069) |
| Cortical volume |  |  |  |  |  |
| SIENAX | **-0.636**<br>**(< 0.001)** | **0.439**<br>**(0.026)** | **-0.557**<br>**(0.009)** | **0.475**<br>**(0.014)** | **-0.485**<br>**(0.011)** |
| recon-all-clinical (T1) | -0.183<br>(0.331) | **0.386**<br>**(0.043)** | -0.293<br>(0.131) | **0.484**<br>**(0.013)** | **-0.369**<br>**(0.048)** |
| recon-all-clinical (FL) | -0.338<br>(0.083) | 0.253<br>(0.189) | -0.255<br>(0.185) | 0.093<br>(0.637) | **-0.392**<br>**(0.038)** |
| WMH-SynthSeg (T1) | -0.098<br>(0.611) | 0.284<br>(0.141) | -0.245<br>(0.198) | 0.384<br>(0.050) | -0.221<br>(0.246) |
| WMH-SynthSeg (FL) | -0.050<br>(0.775) | 0.194<br>(0.292) | -0.157<br>(0.428) | 0.177<br>(0.376) | -0.069<br>(0.699) |
| Thalamic volume<sup>#</sup> |  |  |  |  |  |
| recon-all-clinical (T1) | **-0.476**<br>**(0.029)** | **0.408**<br>**(0.043)** | **-0.433**<br>**(0.033)** | **0.675**<br>**(< 0.001)** | **-0.670**<br>**(0.002)** |
| recon-all-clinical (FL) | **-0.623**<br>**(0.004)** | 0.351<br>(0.111) | **-0.444**<br>**(0.037)** | 0.300<br>(0.185) | **-0.628**<br>**(0.004)** |
| WMH-SynthSeg (T1) | -0.263<br>(0.168) | 0.231<br>(0.231) | -0.331<br>(0.084) | **0.628**<br>**(0.001)** | **-0.410**<br>**(0.029)** |
| WMH-SynthSeg (FL) | -0.250<br>(0.187) | 0.223<br>(0.242) | -0.291<br>(0.131) | **0.407**<br>**(0.040)** | -0.292<br>(0.114) |
| White matter volume |  |  |  |  |  |
| SIENAX | **-0.489**<br>**(0.012)** | 0.306<br>(0.111) | **-0.422**<br>**(0.029)** | **0.414**<br>**(0.039)** | -0.333<br>(0.071) |
| recon-all-clinical (T1) | **-0.393**<br>**(0.044)** | **0.396**<br>**(0.041)** | **-0.428**<br>**(0.029)** | 0.578<br>(0.003) | **-0.531**<br>**(0.005)** |
| recon-all-clinical (FL) | -0.294<br>(0.125) | 0.212<br>(0.260) | -0.201<br>(0.299) | 0.265<br>(0.185) | -0.329<br>(0.074) |
| WMH-SynthSeg (T1) | -0.361<br>(0.061) | **0.402**<br>**(0.040)** | **-0.419**<br>**(0.029)** | **0.614**<br>**(0.001)** | **-0.500**<br>**(0.009)** |
| WMH-SynthSeg (FL) | **-0.432**<br>**(0.033)** | **0.421**<br>**(0.030)** | **-0.425**<br>**(0.029)** | **0.556**<br>**(0.004)** | **-0.500**<br>**(0.009)** |

| Ventricular volume |  |  |  |  |  |
| --- | --- | --- | --- | --- | --- |
| SIENAX | **0.398**<br>(0.044) | **-0.503**<br>(0.020) | **0.500**<br>(0.014) | **-0.558**<br>(0.004) | **0.497**<br>(0.009) |
| recon-all-clinical (T1) | 0.292<br>(0.125) | **-0.378**<br>(0.043) | **0.374**<br>(0.048) | -0.343<br>(0.085) | **0.387**<br>(0.038) |
| recon-all-clinical (FL) | 0.312<br>(0.108) | **-0.482**<br>(0.022) | **0.478**<br>(0.018) | **-0.486**<br>(0.013) | **0.561**<br>(0.004) |
| WMH-SynthSeg (T1) | 0.355<br>(0.062) | **-0.443**<br>(0.026) | **0.422**<br>(0.029) | **-0.400**<br>(0.043) | **0.410**<br>(0.029) |
| WMH-SynthSeg (FL) | 0.322<br>(0.093) | **-0.391**<br>(0.041) | **0.387**<br>(0.042) | -0.383<br>(0.050) | **0.387**<br>(0.038) |

**Legend:** EDSS-Expanded Disability Status Scale; SNRS-Scripts Neurological Rating Scale; Combi-WISE-Combinatorial Weight-Adjusted Disability Score; SDMT- Symbol Digit Modalities Test; ATOPS-Auditory Test of Processing Speed; T1 – T1-weighted ; FL – T2-weighted fluid attenuated inversion recovery

### Bland-Altman plots

Bland-Altman plots for the thalamus and cortex are shown in Figures 2 and 3, respectively, while other tissue segmentations are shown in the Supplemental Figure. Inspection of the plots demonstrated generally good agreement between methods with minimal bias and narrow limits of agreement. However, bias was evident in some comparisons (e.g., recon-all-clinical FLAIR versus recon-all-clinical T1 for cortical GM volume) and many plots had at least one outlier (i.e., outside of the ± 1.96 standard deviation limit of agreement)

**Figure 2.**
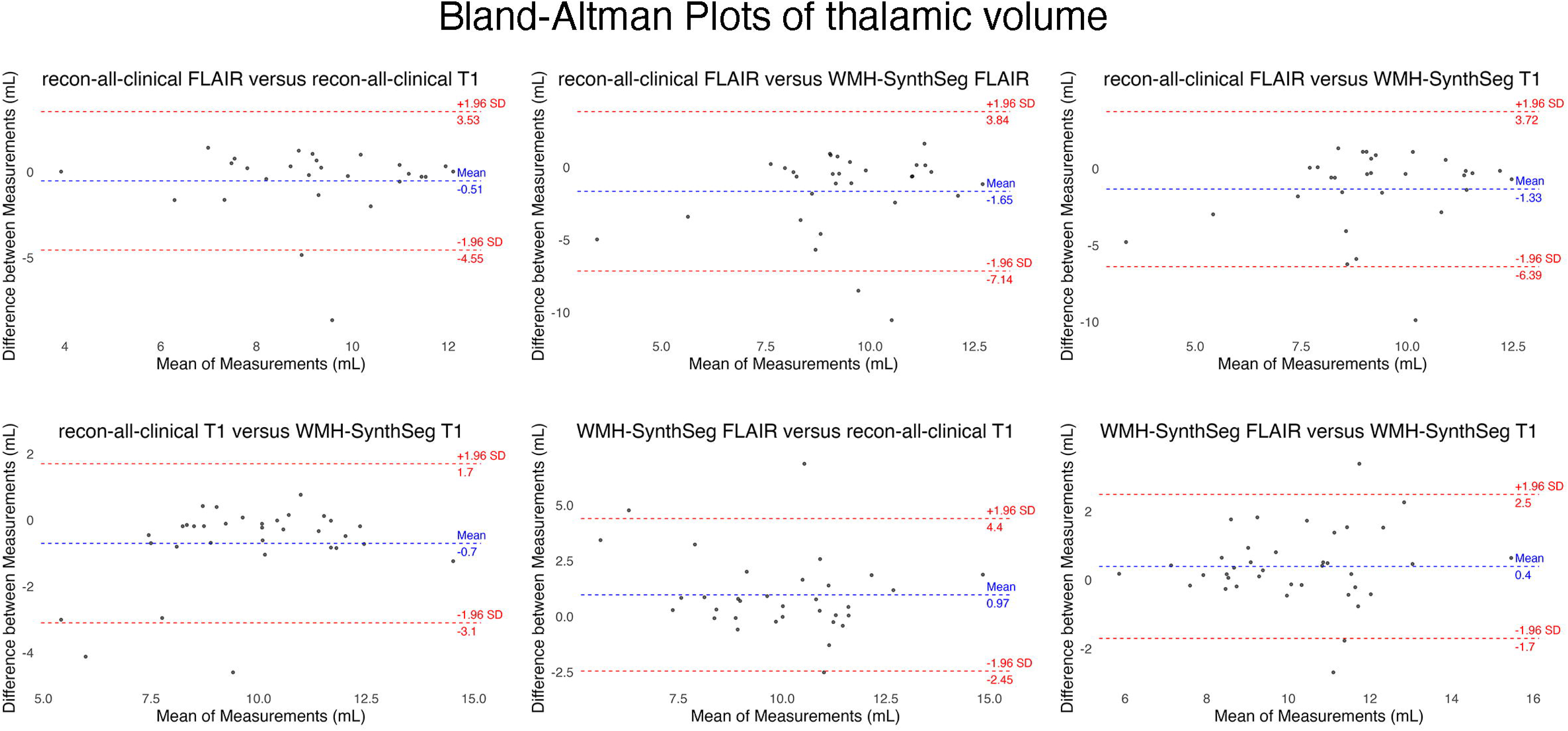
Bland-Altman plots showing comparisons between the different methods for calculating thalamic volume. The blue line indicates the mean difference while the red lines indicate the limits of agreement, defined as the mean difference ± 1.96 standard deviations (SD).

**Figure 3.**
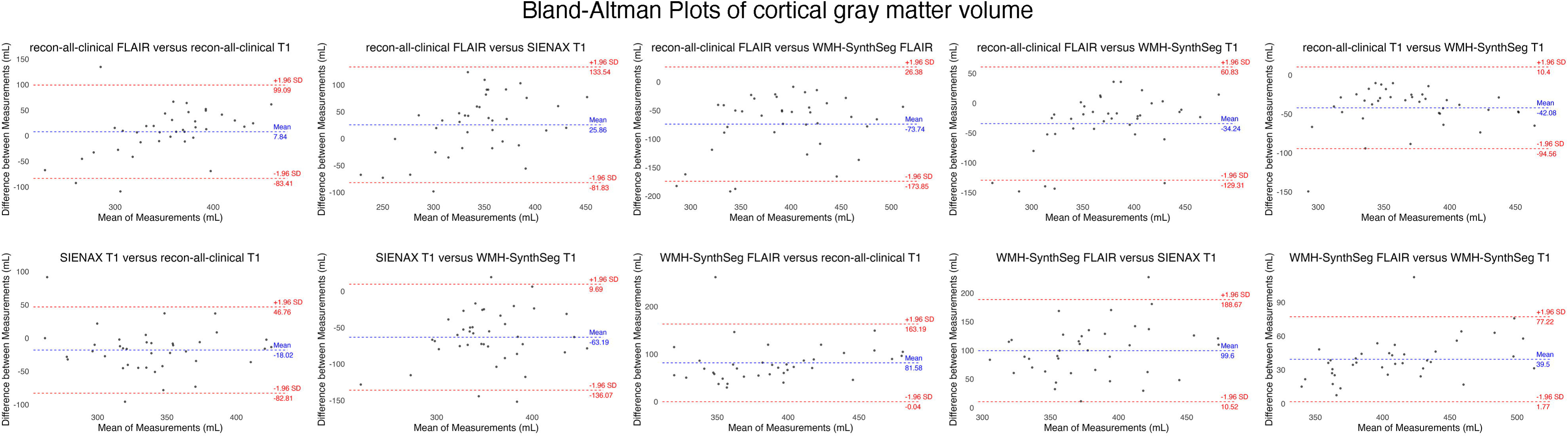
Bland-Altman plots showing comparisons between the different methods for calculating cortical gray matter volume. The blue line indicates the mean difference while the red lines indicate the limits of agreement, defined as the mean difference ± 1.96 standard deviations (SD).

## Discussion

A recent international panel recognized that clinical trial designs, especially in severe MS, need to be more novel, using alternative approaches that would reduce sample size, and prioritize development and application of intermediate outcomes and new technologies in under-researched MS populations.^37^ Because of substantial logistical and conceptual barriers, there is a pressing need for applying and validating novel technologies that can accommodate the complexities of studying individuals affected by severe MS within their living environments.

The findings from this study support our preliminary data,^4^ showing significantly lower brain volumes, particularly for the GM, in those affected by severe MS compared to those with a milder form of the disease. Interestingly, the largest effect sizes were noted for volumes obtained from the SIENAX pipeline, which yields a tissue segmentation based purely on image intensity along with registration-based masks to isolate the cortex and ventricles. Furthermore, cortical GM volume differences were only evidenced with SIENAX; the AI-based methods all had small effect sizes except for recon-all-clinical with the FLAIR as input, which had a medium effect seize albeit not significant. In addition, significant WM volume differences were only detected with the AI-based methods recon-all-clinical (T1 only) and WMH-SynthSeg (both T1 and FLAIR). While we do not have MRI acquisitions from a conventional MRI system (i.e., 1.5T or 3T) to validate our findings, GM pathology is well known to be more strongly associated with increased disability and more advanced disease compared to that in the WM, ^38–41^ suggesting that our SIENAX-derived results have face validity.

We did not detect significant differences between lesion burden and brain volumes in severe MS when comparing early versus late accrual of disability. These findings may suggest that despite initial differences in demographic characteristics and clinical manifestations, early and late onset severe phenotypes, ultimately converge on a common pathophysiological substrate characterized by GM-based neurodegeneration, highlighting the importance of early and continuing neurodegeneration. Future studies should investigate whether those with early accrual of disability have a similar rate of cortical GM atrophy compared to those with late, exploring the hypothesis that MS does not burn out over time in severe MS. A likely cause of GM atrophy in early disability accrual severe MS is primary GM damage (GM lesions),^38^ whereas GM tissue loss in late severe MS may be the combination of primary^38^ and secondary damage (WM lesions leading to secondary retrograde Wallerian degeneration).^42^ A unified GM-centric neurodegenerative process for early and late severe MS may have profound implications for the development of novel therapeutic approaches.

In the CASA-MS phase 1 study, we found that dynamic disability measures (SNRS and Combi-WISE) were more robustly related to MRI outcomes in severe MS compared to the commonly used EDSS.^2^ In the current study, using the entire study population of severe and less severe pwMS, we showed similar associations between increased disability and lower volumes of total GM, cortex (SIENAX only), thalamic (only feasible with AI-driven methods), and WM (AI-driven methods only). We also previously characterized cognitive performance in severe and less severe MS using a newly developed and validated auditory cognitive test (ATOPS), thus eliminating the visual acuity, visual-motor, and memory confounds for which standard testing is criticized in the MS literature.^3^ In line with previous results,^3^ a significantly greater percentage of severe MS completed ATOPS than the current gold standard of SDMT (95.8% vs 79.2%). All in all, these findings suggest that ultra-low-field MRI brain volumetry is useful for investigating clinico-radiological associations in MS.

A comment is also warranted on the associations between tissue volumes and clinical outcomes. All SIENAX-derived volumes were consistently associated with clinical outcomes except for two measures with respect to WM volume; the largest effect was for GM volume and EDSS (r = -0.722). On the other hand, decreased WM volume, which is less reliably associated with disability in MS,^41^ was more consistently found using AI-driven techniques. Taken together with our findings in severe versus less severe MS, our results suggest that data obtained from the recon-all-clinical and WMH-SynthSeg pipelines may be biased to a certain extent and may not fully reflect the true nature of the underlying pathology in the case of MS. Although a recent study validated WMH-SynthSeg for evaluation of Alzheimer’s disease in individuals acquired with ultralow-field portable MRI,^43^ it is conceivable that more subtle differences in cortical atrophy are lost when studying variability across different MS phenotypes. In addition, our imaging protocol did not use isotropic voxels, which may have had an influence on the AI-derived cortical segmentations; it has been shown that hippocampal volumes obtained from ≤ 3mm isotropic ultralow-field MRI using these methods are more accurate than images obtained with anisotropic voxels.^43^ Regardless, the lack of cross-validation of our results with 3T MRI outcomes represents one of the key limitations of the current study. We also lack longitudinal data acquired on the same scanner. Our group is currently working to address both of these issues.

In conclusion, the findings from the CASA-MS phase 2 study demonstrate the feasibility of using ultra-low-field portable MRI in studying severe MS. Moreover, associations were observed between ultralow-field MRI brain volumetry and clinical endpoints across a broad spectrum of disease severity, suggesting its potential for assessing neurodegeneration and disability progression in those individuals living with severe MS. However, careful consideration is required in implementing tissue volumetry pipelines as findings are heavily dependent on the choice of algorithm and input.

## Acknowledgements

We express our profound gratitude to the patients and caregivers whose invaluable contributions have been the cornerstone of this research, offering unique insights and experiences vital to our study on severe MS. Their resilience and dedication have been inspirational. We also extend our heartfelt thanks to our fellow researchers for their expertise and unwavering commitment to scientific excellence. This collaborative effort between the Boston Home and the University at Buffalo underscores the importance of integrating real-world experiences with rigorous academic research to advance our understanding of conditions, like is severe MS.

---

**Supplemental Figure.**
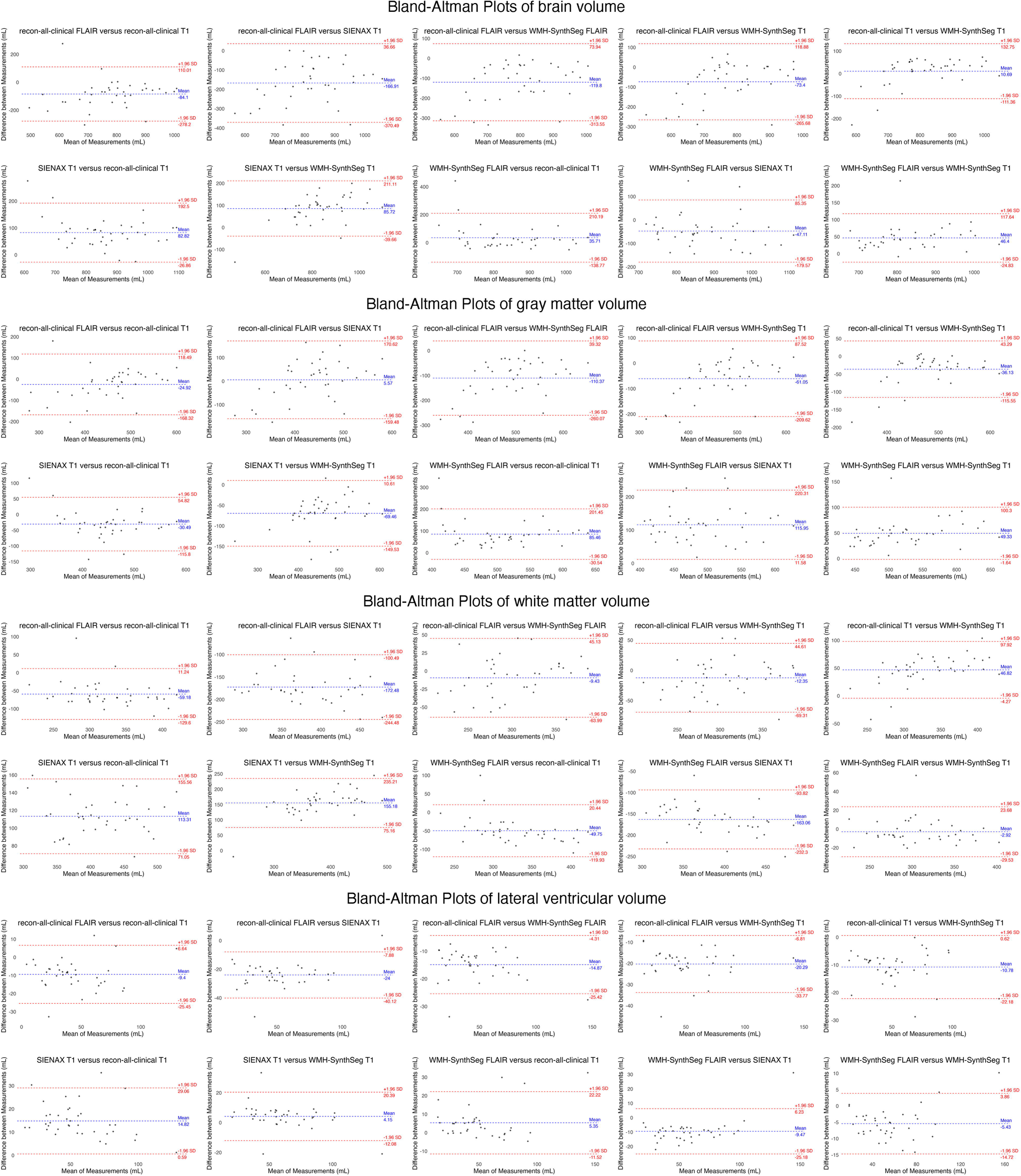
Bland-Altman plots showing comparisons between the different methods for calculating brain volume, gray matter volume, white matter volume, and lateral ventricular volume. The blue line indicates the mean difference while the red lines indicate the limits of agreement, defined as the mean difference ± 1.96 standard deviations (SD).

